# Prevalence and associated factors of burnout in allied healthcare professionals in a tertiary hospital in Singapore

**DOI:** 10.1101/2020.08.16.20176206

**Authors:** Yao Hao Teo, Jordan Thet Ke Xu, Cowan Ho, Jui Min Leong, Benjamin Kye Jyn Tan, Elisabeth Ker Hsuen Tan, Wei-An Goh, Elson Neo, Jonathan Yu Jing Chua, Sean Jun Yi Ng, Julia Jie Yi Cheong, Jeff Yi-Fu Hwang, See Ming Lim, Thomas Soo, Judy Gek Khim Sng, Siyan Yi

**Affiliations:** Yong Loo Lin School of Medicine, National University of Singapore, Singapore, Singapore; Saw Swee Hock School of Public Health, National University of Singapore and National University Health System, Singapore, Singapore; Occupational Health Clinic, Department of Medicine, National University Hospital, National University Health System, Singapore, Singapore; KHANA Center for Population Health Research, Phnom Penh, Cambodia; Center for Global Health Research, Touro University California, Vallejo, CA, USA; School of Public Health, National Institute of Public Health, Phnom Penh, Cambodia

**Keywords:** Mental health, Healthcare provider, Occupational health, Work environment, psychological support, Asia

## Abstract

**Objective:** To explore the prevalence of burnout, factors associated with burnout, and barriers to seeking psychological help among allied health professionals in Singapore.

**Design:** Cross-sectional study.

**Setting:** A tertiary hospital in Singapore.

**Participants:** Allied health professionals.

**Primary outcome measure:** Burnout measured by using Maslach Burnout Inventory (MBI-HSS)

**Results:** In total, 328 participants completed the questionnaire. The prevalence of burnout was 67.4%. A majority of the respondents were female (83.9%), Singaporean (73.5%), aged 40 years and below (84.2%), and of Chinese ethnicity (79.9%). In the multiple logistic regression model, burnout was negatively associated with being in the age groups of 31 to 40 (AOR 0.39, 95% CI 0.16–0.93) and ≥40 years (AOR 0.30, 95% CI 0.10–0.87) and a low workload burden (AOR 0.35, 95% CI 0.23–0.52). Burnout was positively associated with a longer work experience of 3 to 5 years (AOR 5.27, 95% CI 1.44–20.93) and more than five years (AOR 4.24; 95% CI 1.16–16.79). Among the 190 participants who completed the PBPT, barriers to seeking psychological help shown to be significantly associated with burnout were a ‘lack of motivation’ and ‘time constraints’ (p< 0.01).

**Conclusions:** This study shows a high prevalence of burnout and identifies its associated factors among Singapore’s allied health professionals. The findings revealed the urgency of addressing burnout in allied health professionals and the need for effective interventions to reduce burnout. Concurrently, proper consideration of the barriers to seeking help is warranted to improve allied health professionals’ mental wellbeing.

**Strengths and limitations of this study:**

- This study is the first to show a high prevalence of burnout and identify its associated
- We used validated scales, including Maslach Burnout Inventory, Areas of Worklife Survey, and Perceived Barriers to Psychological Treatment.
- Limitations of the study included the cross-sectional design, the low response rate, the potential social desirability bias, and the limited generalizability.
- This study did not cover factors associated with burnout in previous studies, including the increasing computerization of practices and the participants’ personality traits.

## INTRODUCTION

Burnout is a prolonged response to chronic emotional and interpersonal stressors on the job, comprising three dimensions: exhaustion, cynicism, and inefficiency.^1^ These dimensions are further defined as follows: exhaustion of emotional or physical capacity due to stress, a degree of indifference or detachment from various aspects of work, and a sense of inadequacy or reduced personal accomplishment, respectively.^1^

In healthcare settings, burnout negatively impacts outcomes at the individual, interpersonal, and institutional levels. At the individual level, burnout is associated with reduced job satisfaction, increased absenteeism, medical errors, sickness, injury, and accidents among healthcare providers.^2 3^ These individual-level impacts may lead to reduced care quality and higher mortality levels among patients.^4 5^ From an interpersonal perspective, burnout is known to be associated with emotional dissonance due to chronic exhaustion and cynicism.^6^ Emotional dissonance is described as a conflict between personal emotions and organizational demands. On an institutional level, burnout is linked to a higher turnover of healthcare workers,^7 8^ and decreased workforce efficiency,^9^ posing a substantial economic burden on the healthcare system.^10^

The pernicious nature of burnout in healthcare settings has prompted numerous studies on its prevalence in Singapore and internationally. For example, high rates of burnout and its associations among physicians and nurses have been reported in Singapore.^11 12^ Extensive research involves the barriers to seeking help for doctors, such as fear of stigma, lack of available time, and lack of convenient access.^13 14^

Allied health professionals (AHPs) are defined as individuals who work collaboratively to deliver health or related services distinct from medicine or nursing,^15^ covering frontline staff involved in the provision of routine and essential services in medical care.^16^ AHPs include, but are not limited to, occupational therapists, physiotherapists, pharmacists, medical social workers, and medical technologists.^17^ In Singapore, the Allied Health Professions Council (AHPC) further classified allied health occupations.^18^

Studies in other countries have reported a high prevalence of burnout in AHPs. In the United States, physiotherapists and occupational therapists reported high rates of emotional exhaustion (58%), negative feelings about their work and their clients (94%) as well as an almost non-existent sense of personal accomplishment (1%).^19^ However, there are currently no studies examining the prevalence and associations of burnout among AHPs in Singapore.

Hence, this study aims to identify the prevalence of burnout and explore associations of sociodemographic factors and work environment with burnout among AHPs in Singapore. Our secondary objective is to identify significant barriers in seeking psychological help among AHPs.

## METHODS

### Study design and sampling

We conducted a cross-sectional study among AHPs working in a tertiary acute care hospital between October 2019 to December 2019. Based on previous studies looking at the prevalence of burnout in AHPs and the total number of AHPs in Singapore,^20 21^ we determined the sample size through the application of a single proportion formula with the assumption of 60% prevalence, 5% marginal error, and 95% confidence level (CI). The minimum required sample size for the study was 348.

### Inclusion and exclusion criteria

Staff members in the tertiary hospital who were classified as an AHP based on the definition established by the AHPC in Singapore were eligible to participate in the study.^18^ They included AHPs of all seniority levels, including frontline staff who are described as healthcare workers involved in the provision of routine and essential services in medical care.^16^

### Questionnaire design and measurement

We developed an electronic survey and emailed all AHP staff working for the tertiary hospital to request their participation. The survey comprised four components: (1) sociodemographic characteristics, (2) Maslach Burnout Inventory (MBI-HSS), (3) Areas of Worklife Survey (AWS), and (4) Perceived Barriers to Psychological Treatment (PBPT).

Sociodemographics questions were adapted from the Singapore National Health Survey 2010,^22^ covering residency status, age, gender, ethnicity, income levels, caregiver status, occupation, employment history, physical activity levels, and mental health.

We assessed burnout by using the Maslach Burnout Inventory (MBI), in particular, the MBI-Human Services Survey for Medical Personnel MBI-HSS(MP).^23^ MBI has been widely used in different settings.^24^ The 22-item standardized questionnaire assessed three subscales based on the definition of burnout. These are emotional exhaustion (EE), depersonalization (DP), and personal accomplishment (PA). It is a validated tool with a Cronbach’s alpha of 0.90 for EE, 0.79 for DP, and 0.71 for PA.^25^ Among the three subscales, PA was excluded from this study because its association with burnout has been more variable and complex, similar to previous studies.^26^ Hence, we defined burnout as experiences of a high level of EE, DP, or both.^27^ While there is no universal cut-off score for the MBI, this study defined high EE as a cut-off score of more than or equal to 27 points, and high DP as a cut-off score of more than or equivalent to 10 points, based on cut-offs used in previous studies.^28 29^

The AWS is a 28-item scale that is part of the MBI toolkit.^30^ The scale examines the dimensions of an individual’s work life and predicts their relationship with burnout.^31^ It has been widely used^32 33^ and validated (Cronbach’s alpha: 0.71 to 0.85)^31^ in healthcare settings. The six dimensions assessed in the survey were: workload, control, reward, community, fairness, and values. They are defined as follows: “Workload” refers to the employee’s ability to cope with work demands. “Control” refers to the level of active involvement of an employee in work decisions. “Reward” refers to rewards that place higher value and recognition on an employee’s work. “Community” refers to the overall quality of social interaction at work. “Fairness” refers to the general equity of decisions made at the workplace. And “Values” refers to the dissonance between personal and organizational values.^34^ A higher AWS score indicates a more balanced relationship, rather than conflicted one,^31^ between the respondent and their work.^35^

The last component of the survey comprised the 27-item PBPT questionnaire, which has been previously validated (Cronbach’s alpha: 0.92).^36^ Items are classified into nine domains: stigma, lack of motivation, emotional concerns, negative evaluations of therapy, misfit of therapy to needs, time constraints, participation restriction, availability of services, and cost.^36^ We asked participants to rate on a 5-point Likert scale the degree to which each item hindered them from seeing a counselor or a therapist. A score of 4–5 was deemed as “substantial barriers.” A domain is deemed to represent a “substantial barrier” if at least one item within that domain was reflected as a “substantial” barrier.” Given the lengthy questionnaire and to improve the overall response rate and sample size,^37^ we made the PBPT questionnaire component optional for participants in this study.

### Data analyses

We used R Commander version 2.7.11 to perform all statistical analyses. We computed Cronbach’s alpha for each MBI subscale and AWS domain to assess reliability. We performed Bivariate analyses of the demographic factors and the AWS dimensions to examine their association with burnout using Pearson’s Chi-square test or Fisher’s exact tests (when a cell count was smaller than five). We identify factors associated with burnout by using multiple logistic regression analysis. We entered variables with statistical significance (p< 0.05) in bivariate analyses simultaneously in the multiple logistic regression model. For respondents who completed the optional component on PBPT, we recorded the incidence of expressing a variable as a “substantial barrier.” We compared it between the burnout and non-burnout groups using Pearson’s Chi-squared test.

### Ethical considerations

The National Healthcare Group Domain Specific Review Board approved this study(2019/00477). No identifiable information of participants was collected. We stored all data on a secure, Health Insurance Portability, and Accountability Act compliant, web-based server (REDCap). We included a participant information sheet in the email, providing all relevant information on participant anonymity and consent for voluntary participation.

## RESULTS

### Sociodemographic characteristics

Among the 1127 eligible AHPs invited, 345 participated in the survey. However, we excluded 17 questionnaires due to incomplete entries. We included a total of 328 respondents in the analyses, providing a response rate of 29.1%.

Table 1 shows the sociodemographic characteristics of the respondents. A majority of the respondents were Singaporean (73.5%), aged 40 years and below (84.2%), female (83.9%), and of Chinese ethnicity (79.9%). Almost all respondents were working full time(94.2%). Among the occupational groups, dieticians (94.7%) and pharmacists (82.5%) had the highest burnout prevalence. More than half of the respondents had worked for more than five years in the same organization. Approximately half of the respondents worked as frontline staff and reported low levels of physical activity. Only a small proportion of the respondents reported a history of mental illness or had sought help from a professional within the past year for mental illness.

**Table 1.** Demographics of allied health professionals in the study stratified by burnout status

| Variables | Number<br>(*n*= 328)<br>*n* (%) | With burnout<br>(*n*= 221)<br>*n* (%) | Without burnout<br>(*n*= 107)<br>*n* (%) | *p*-Value |
| --- | --- | --- | --- | --- |
| Residency status |  |  |  |  |
| Singaporean | 241 (73.5) | 164 (68.0) | 77 (32.0) | 0.10 |
| Age group |  |  |  |  |
| 21 to 30 | 137 (41.8) | 101 (73.7) | 36 (26.3) |  |
| 31 to 40 | 139 (42.4) | 94 (67.6) | 45 (32.4) |  |
| 41 years and above | 52 (15.9) | 26 (50.0) | 26 (50.0) |  |
| Male gender | 53 (16.1) | 36 (67.9) | 17 (32.1) | 1.00 |
| Chinese ethnic group | 262 (79.9) | 182 (69.5) | 80 (30.5) | 0.14 |
| Average monthly household income |  |  |  | 0.14 |
| Less than S\$5000 | 25 (7.6) | 18 (72.0) | 7 (28.0) | |
| S\$5000 to \$9000 | 118 (36.0) | 85 (72.0) | 33 (28.0) | |
| S\$9000 and above | 109 (33.2) | 64 (58.7) | 45 (41.3) | |
| Not disclosed | 76 (23.2) | 54 (71.1) | 22 (28.9) |  |
| Caregiver status - Yes | 72 (22.0) | 48 (66.7) | 24 (33.3) | 0.96 |
| Occupation |  |  |  | 0.07 |
| Clinical psychologist | 5 (1.5) | 2 (40.0) | 3 (60.0) |  |
| Radiographer | 32 (9.8) | 19 (59.4) | 13 (40.6) |  |
| Dietician | 19 (5.8) | 18 (94.7) | 1 (5.3) |  |
| Medical Technologist | 96 (29.3) | 64 (66.7) | 32 (33.3) |  |
| Medical Social Worker | 19 (5.8) | 15 (78.9) | 4 (21.1) |  |
| Occupational Therapist | 33 (10.0) | 21 (63.6) | 12 (36.4) |  |
| Pharmacist | 40 (12.2) | 33 (82.5) | 7 (17.5) |  |
| Physiotherapist | 28 (8.5) | 17 (60.7) | 11 (39.3) |  |
| Podiatrist | 5 (1.5) | 3 (60.0) | 2 (40.0) |  |
| Respiratory Therapist | 5 (1.5) | 2 (40.0) | 3 (60.0) |  |
| Speech Therapist | 17 (5.2) | 10 (58.8) | 7 (41.2) |  |
| Others | 29 (8.8) | 17 (58.6) | 12 (41.4) |  |
| Duration working at the current organization |  |  |  | <0.01 |
| <1 year | 24 (7.3) | 9 (37.5) | 15 (62.5) |  |
| 1-2 years | 47 (14.3) | 31 (66.0) | 16 (34.0) |  |
| 3-5 years | 64 (19.5) | 53 (82.8) | 11 (17.2) |  |
| >5 years | 193 (58.8) | 128 (66.3) | 65 (33.7) |  |
| Being frontline staff | 175 (53.4) | 116 (66.3) | 59 (33.7) | 0.98 |
| Full-time employed | 309 (94.2) | 214 (69.3) | 95 (30.7) | <0.01 |
| Average number of night shifts per month |  |  |  | 0.24 |
| 1-3 times | 33 (10.0) | 25 (75.8) | 8 (24.2) |  |
| 4-6 times | 18 (5.5) | 12 (66.7) | 6 (33.3) |  |
| 7 or more times | 4 (1.2) | 1 (25.0) | 3 (75.0) |  |
| Not applicable | 273 (83.2) | 183 (67.0) | 90 (33.0) |  |
| Low level of physical activity | 185 (56.6) | 129 (69.7) | 56 (30.3) | 0.58 |
| History of mental illness | 7 (2.2) | 6 (85.7) | 1 (14.3) | 0.43 |
| Sough medical help in the past year | 15 (4.7) | 15 (100.0) | 0 (0.0) | <0.01 |

The Cronbach’s alpha coefficients for EE, DP, and PA in MBI-HSS were 0.93, 0.81, and 0.85, respectively, suggesting that the overall measurement was reliable. As shown in Table 1, full-time workers were significantly more likely to experience burnout than part-time workers. Respondents with more than one year of work experience reported a higher prevalence of burnout than those with less than one year of work experience. Respondents who had sought professional mental help in the past year were significantly more likely to have burnout than those who did not.

#### Burnout prevalence

The prevalence of burnout among AHPs in this study was 67.4%. Figure 1 demonstrates the numbers of AHPs with a burnout in each subdomain among the AHPs. A majority of the respondents reported high levels of burnout on EE, less than half reported a high level on DP, and more than one-third had both high EE and DP. Despite a large proportion of AHPs in this study having high EE or DP, most of them also had high scores for PA.

**Figure 1.**
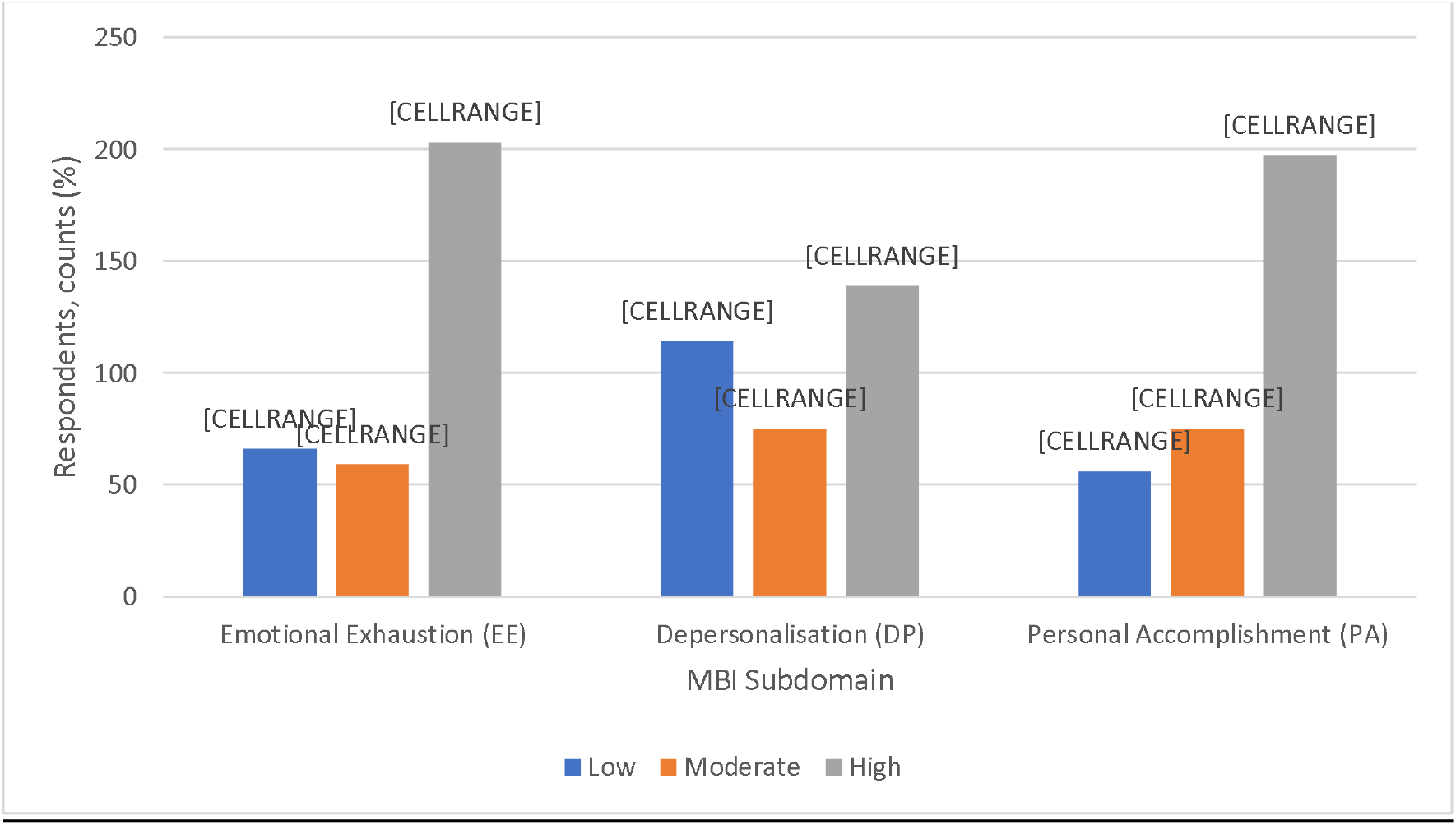
Frequency of allied health professionals by degree (low, moderate, high) in each subdomain of burnout (*n* = 328)

#### AWS domains and association with burnout

The Cronbach’s alpha coefficients for workload, control, reward, community, fairness, and values were 0.78, 0.77, 0.89, 0.86, 0.82, and 0.78, respectively. As shown in Figure 2, all domains of AWS were significantly associated with burnout (p≤0.01), with workload, control, and reward showing the most significant differences in the mean scores between burnout and non-burnout groups.

**Figure 2.**
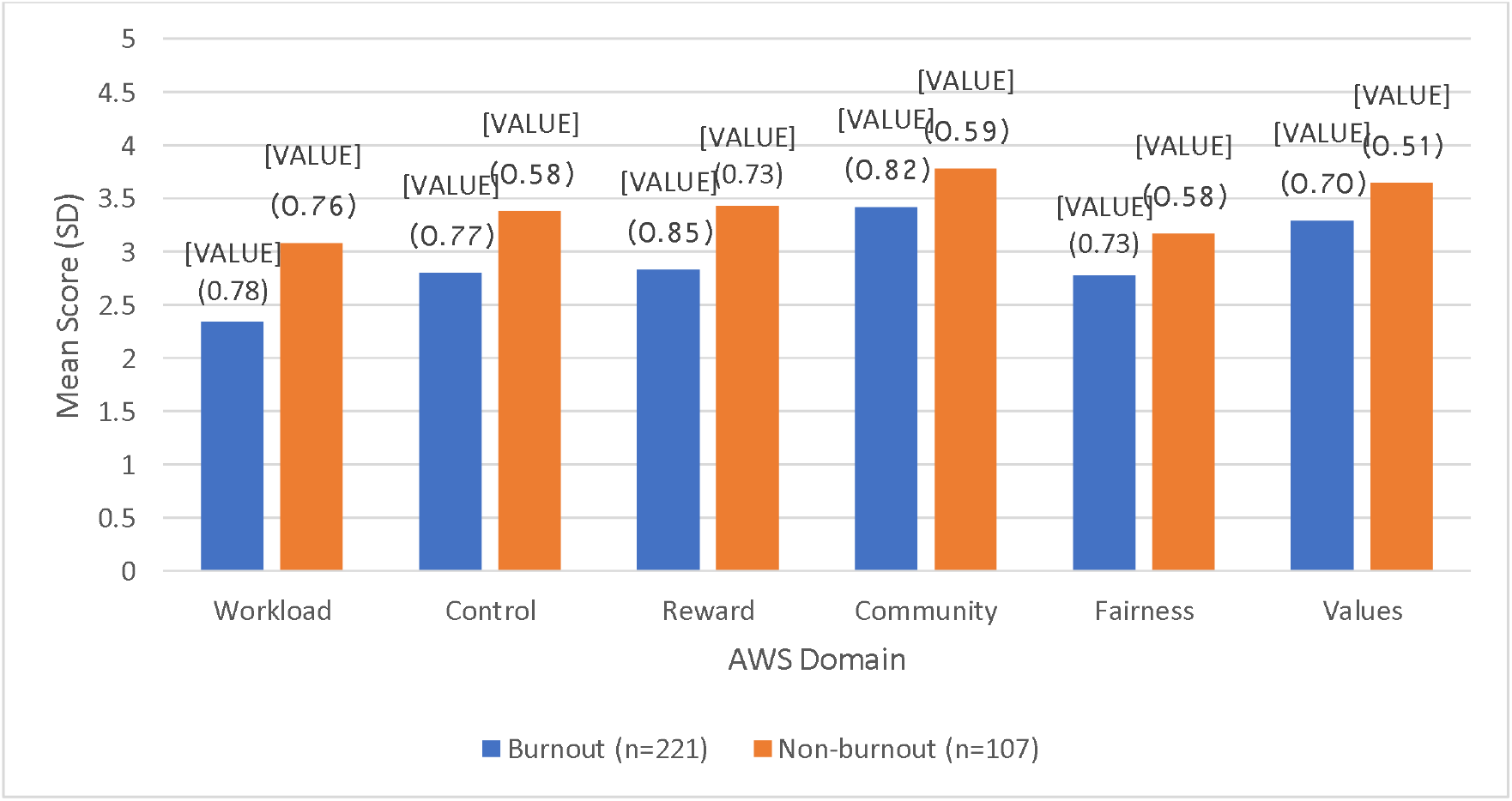
Comparisons of the mean scores of the Areas of Worklife Survey domains stratified by burnout (*n* = 328)

#### AWS domain and association with burnout

Figure 3 presents the absolute mean score differences of responses to individual AWS statements between burnout and non-burnout groups (see Table 1a in the Appendix for detailed results). The majority of the mean score differences in all domains were significant. The workload domain had the highest absolute difference compared to the other domains. The statements “I have so much work to do on the job that it takes me away from my personal interests” and “I do not have time to do the work that must be done” in the workload domain scored the highest absolute difference in mean scores among all questions.

**Figure 3.**
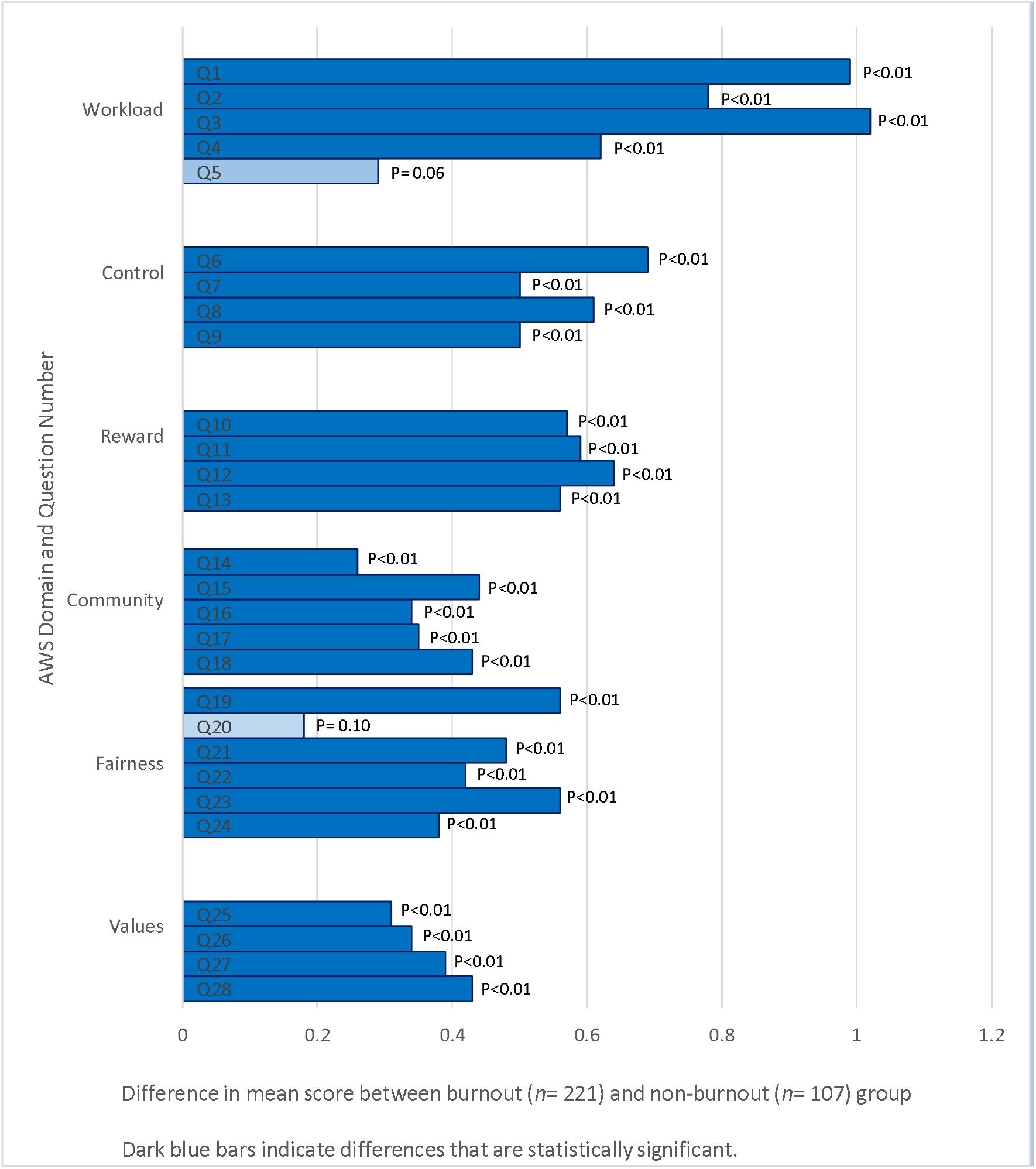
Comparisons of difference in mean scores of Areas of Worklife Survey statements in all domains between burnout (*n* = 221) and non-burnout (*n* = 107) group

#### Factors associated with burnout

In the multiple logistic regression model (Table 2), AHPs who had lower mean scores in the workload subdomain of the AWS, indicative of a high workload burden, were almost three times more likely to be burnout than those who had higher mean scores. Compared to respondents aged 30 years and below, older AHPs aged 31 and above were significantly less to have burnout. Moreover, respondents who had worked in the current organization for more than three years were approximately five times more likely to experience burnout than respondents who had worked in the current organization for less than one year.

**Table 2.** Factors associated with burnout in multiple logistic regression analysis

|  | Coefficient (SE) | AOR (95% CI) | P-value |
| --- | --- | --- | --- |
| AWS domain |  |  |  |
| Workload | -1.05 (0.21) | 0.35 (0.23, 0.52) | <0.01 |
| Control | -0.46 (0.29) | 0.63 (0.35, 1.10) | 0.11 |
| Reward | -0.34 (0.25) | 0.71 (0.43, 1.17) | 0.18 |
| Community | -0.39 (0.26) | 0.68 (0.40, 1.12) | 0.13 |
| Fairness | -0.18 (0.32) | 0.83 (0.45, 1.54) | 0.56 |
| Values | -0.10 (0.30) | 0.90 (0.50, 1.63) | 0.74 |
| Age group |  |  |  |
| 21 to 30 | Reference | 1.00 | - |
| 31 to 40 | -0.93 (0.45) | 0.39 (0.16, 0.94) | 0.04 |
| 41 years and above | -1.20 (0.55) | 0.30 (0.10, 0.87) | 0.03 |
| Duration working at the current organization |  |  |  |
| <1 year | Reference | 1.00 | - |
| 1-2 years | 0.61 (0.68) | 1.84 (0.50, 7.23) | 0.37 |
| 3-5 years | 1.66 (0.68) | 5.27 (1.44, 20.91) | 0.01 |
| >5 years | 1.44 (0.68) | 4.23 (1.16, 16.76) | 0.03 |
| Employment status |  |  |  |
| Part-time | Reference | 1.00 | - |
| Full-time | 0.62 (0.60) | 1.86 (0.58-6.42) | 0.30 |
| Sough medical help in the past year |  |  |  |
| No | Reference | 1.00 | - |
| Yes | 17.13 (854.15) | 27398446.44 (NA) | 0.98 |

Abbreviations: AWS, Areas of Worklife Survey; SE, standard error; CI, confidence interval; AOR, adjusted odds ratio.

#### Relationship of burnout and perceived barriers to seeking help

As shown in Table 3, 57.9% of participants completed the optional component on PBPT. Among the respondents, barriers to seeking psychological help shown to be significantly associated with burnout were a ‘lack of motivation’ and ‘time constraints’ (p< 0.01).

**Table 3.**
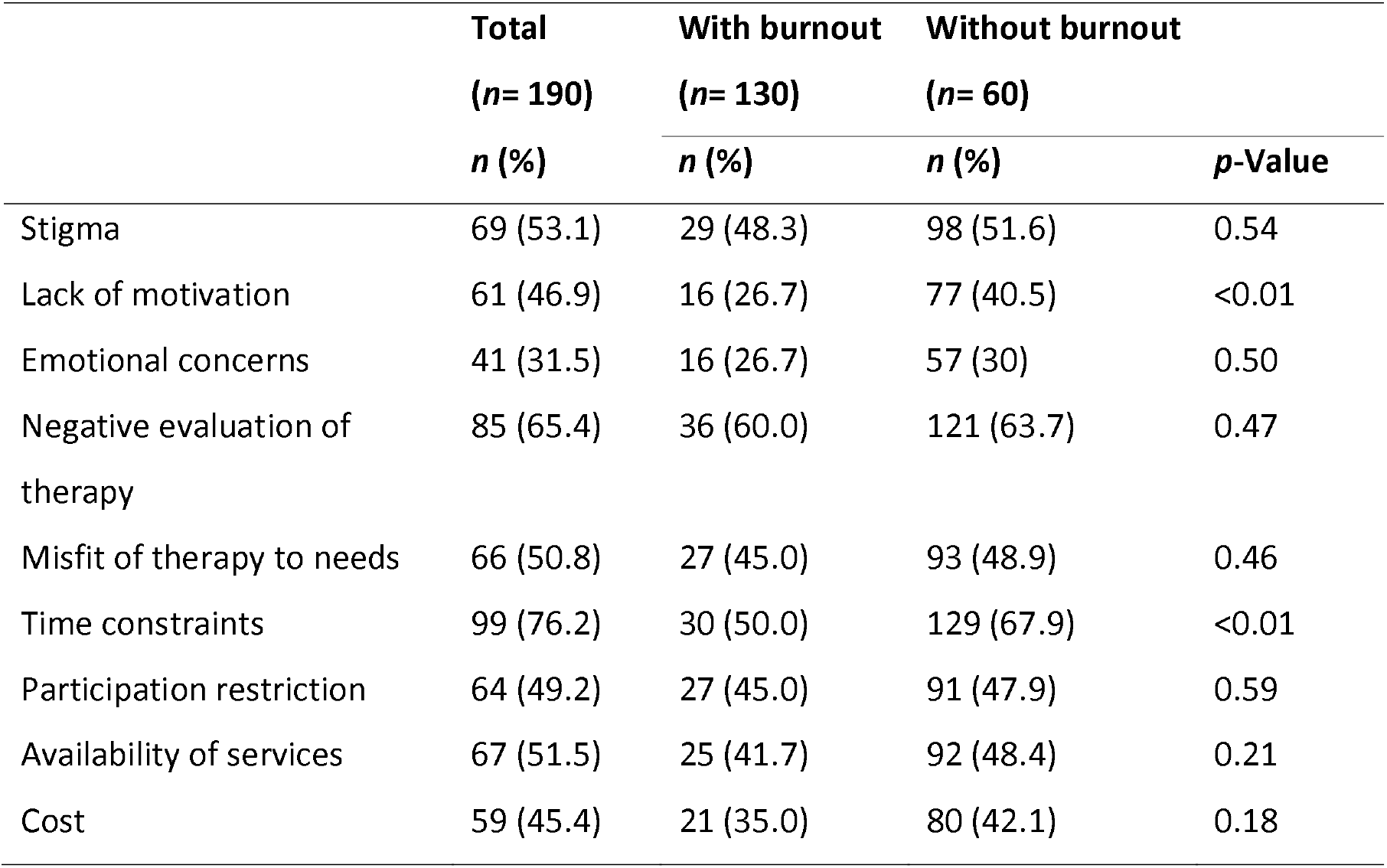
Barriers to seeking psychological help associated with burnout (*n* = 190)

| | Total<br>($n= 190$)<br>*n (%)* | With burnout<br>($n= 130$)<br>*n (%)* | Without burnout<br>($n= 60$)<br>*n (%)* | *p*-Value |
| --- | --- | --- | --- | --- |
| Stigma | 69 (53.1) | 29 (48.3) | 98 (51.6) | 0.54 |
| Lack of motivation | 61 (46.9) | 16 (26.7) | 77 (40.5) | <0.01 |
| Emotional concerns | 41 (31.5) | 16 (26.7) | 57 (30) | 0.50 |
| Negative evaluation of<br>therapy | 85 (65.4) | 36 (60.0) | 121 (63.7) | 0.47 |
| Misfit of therapy to needs | 66 (50.8) | 27 (45.0) | 93 (48.9) | 0.46 |
| Time constraints | 99 (76.2) | 30 (50.0) | 129 (67.9) | <0.01 |
| Participation restriction | 64 (49.2) | 27 (45.0) | 91 (47.9) | 0.59 |
| Availability of services | 67 (51.5) | 25 (41.7) | 92 (48.4) | 0.21 |
| Cost | 59 (45.4) | 21 (35.0) | 80 (42.1) | 0.18 |

## DISCUSSION

This study is the first to investigate the prevalence of burnout and its related factors among AHPs in Singapore. We found a high prevalence of burnout at 67.4% among AHPs in a tertiary hospital. Based on the job demands-resources model of burnout, high EE, and DP scores in our study demonstrates a high probability of resource conservation by AHPs. AHPs may spend less time with patients, resulting in increased clinical errors,^38^ and negatively impacting patient care. However, compared to a study conducted among physical and occupational therapists in the United States, while the EE scores were similar (58% vs. 62% in our study), the DP scores in our study were significantly lower (94% vs. 42% in our study).^19^ The relatively lower prevalence of depersonalization may be attributed to the participants’ organizational factors such as different healthcare systems and different attitudes towards work between AHPs in Asian and Western societies.^39^

Of note, the high prevalence of burnout in pharmacists (82.5%) and dieticians (94.7%) is concerning. We postulate that pharmacists may be prone to experiencing burnout and lower job satisfaction than other occupations with more job variety reported in previous studies.^40^ However, similar studies have shown that dieticians score lower EE than comparison groups of doctors, nurses, and social workers,^20^ indicating lower burnout. Hence, the high prevalence of burnout among dieticians may be due to other organizational or demographic factors. As the sample size of pharmacists and dieticians in this study was small, these associations were not significant. Further studies will be warranted to identify the associated factors of burnout.

In the multiple regression analysis, we found a higher prevalence of burnout in the younger age group of 21 to 30 than in AHPs aged 31 years and above. Previous studies have supported this trend of burnout affecting younger employees.^40 41^ Compared to younger employees, older workers have been better at handling occupational stress and are less prone to burnout.^42^

Work experience may play an essential role in the prevalence of burnout. Employees who have worked for a longer duration (three years and above) in the same organization were more likely to be burnout compared to those who worked for less than a year. We postulate that this could be due to long-term exposure to the patient suffering at the workplace, resulting in emotional exhaustion.^43 44^

We found that heavier workloads contributed to a higher prevalence of burnout among the AHPs. It was the only subdomain of the AWS, significantly associated with burnout after adjusting for covariates. Previous studies have shown the adverse effects of increased workloads among healthcare workers, manifesting burnout.^45 46^ In our study, we demonstrated that this association holds for AHPs in Singapore. In particular, the association of heavier workload among burnout AHPs is most apparent when the workload interferes with their “personal interests” and “work that must be done.”

Hence, the identified associated factors of burnout highlight the need to address potential stressors at work. The intervention may be conducted through both individual and organizational evidence-based strategies.^47^ Given the high EE and DP levels, previous studies have shown the effectiveness of interventions that target personal coping skills such as mindfulness and stress management training, and cognitive-behavioral interventions in reducing occupational stress levels.^48^ Concurrently, given that heavy workload and more extended work experience is associated with burnout, workplace interventions are crucial. A case example will be the United Kingdom-commissioned review.^49^ The review proposes a whole-system workplace intervention, from understanding local staff requirements, multi-level staff engagement, strong visible leadership, support for wellbeing at board level, and a focus on management capability to improve mental wellbeing, lowering burnout.

Lastly, among the subgroup who completed the PBPT questionnaire, ‘lack of motivation’ and ‘time constraints’ were identified as significant barriers to seeking psychological help. Firstly, motivation refers to the general or therapy-focused pursuit of goals,^36^ and a lack of it may be both a symptom and a causative factor of burnout. Regardless of which, motivation was previously reported to be critical in treatment effectiveness,^50^ and further studies examining the role of motivation in seeking help for burnout is crucial before any interventions can be proposed. Secondly, time restraints highlight that the daily responsibilities of AHPs may contribute to burnout and also compete for time, hence a barrier in undergoing therapy. Daily responsibilities include formal duties to their patients and adjunct activities such as documentation, communication, following up on treatment, performing roll calls, or handing over. These auxiliary activities underestimate the time spent on the job.^51^ Hence, accounting for the adjunct activities and enforcing stricter regulations in total work hours may be essential to improve uptake of AHPs in seeking help for their burnout.

There are a few limitations to this study. First, the response rate to the survey was only 29.1%. The response rate was lower than that of healthcare worker surveys in general^52^ and translated to a significant non-response bias for the study. Despite utilizing approaches to increase the response rate, such as through the engagement of respective departmental heads and email reminders, the survey response remained low. The low response rate may lead to a falsely lower burnout rate in our study. Second, burnout is multi-factorial, and this study may not capture the full spectrum of variables. Factors that have not been covered in this study include the increasing computerization of practice,^53^ and the personality traits of the participants.^54^ Third, this study’s cross-sectional nature does not allow the authors to determine causal relationships between the risk factors and burnout. Further longitudinal studies will be needed. Lastly, participant response could have been influenced by social desirability bias due to the highly stigmatized perception of burnout in the workplace.

## CONCLUSION

This study is the first to show a high prevalence of burnout and identify its associated factors among AHPs in Singapore. The prevalence of burnout among AHPs in this study was 67.4%. The identified risk factors of burnout included increased workload, lesser work experience, and younger age. Besides, respondents reported the lack of motivation and time constraints as significant barriers associated with seeking help for burnout. The findings revealed the significance and urgency of addressing burnout in these vulnerable target groups. There is also a potential need to implement individual and organizational interventions such as mindfulness and stress management training, cognitive-behavioral interventions, or workplace interventions that target organizational, cultural, social, and physical aspects of staff health. These interventions should be implemented with proper consideration of the barriers to seeking help to reduce burnout risk effectively. Further longitudinal studies will be useful in exploring the causal relationship between the risk factors and burnout to characterize burnout’s nature better. Concurrently, qualitative studies will be necessary to better understand the broad and complex spectrum of factors influencing burnout among AHPs in Singapore.

## Data Availability

Data used for this study can be accessed upon request from the Principal Investigator (Dr. Siyan Yi) at.

## Acknowledgments

The authors would like to thank the faculty members of Saw Swee Hock School of Public Health and the Yong Loo Lin School of Medicine, whose advice and ideas were integral to this study’s success. The authors would like to thank the anonymous reviewers whose input and feedback significantly improved this manuscript. Lastly, the authors would like to thank the Community Health Project team members for their contributions to the study’s conceptualization.

## Contributors

YHT, TKXJ, CH, BTKJ, ETKH, GWA, EN, and SY designed the study and developed the study protocol and tools. YHT, JH, LSH, TS, and JS were responsible for data collection. YHT, TKXJ, LJM, and SY analyzed data and wrote the manuscript. All authors contributed to the conceptualization of the research questions, interpretation of the results, and manuscript writing. All authors read and approved the final manuscript.

## Ethics approval

The National Healthcare Group Domain Specific Review Board approved the study (2019/00477).

## REFERENCES

1. Maslach C, Schaufeli WB, Leiter MP. Job burnout. Annual review of psychology 2001;52:397–422. doi:10.1146/annurev.psych.52.1.397 [published Online First: 2001/01/10]

2. Silva SC, Nunes MA, Santana VR, et al. Burnout syndrome in professionals of the primary healthcare network in Aracaju, Brazil. Cien Saude Colet 2015;20(10):3011–20. doi:10.1590/1413-812320152010.19912014 [published Online First: 2015/10/16]

3. Bartosiewicz A, Januszewicz P. Readiness of Polish Nurses for Prescribing and the Level of Professional Burnout. Int J Environ Res Public Health 2018;16(1):35. doi:10.3390/ijerph16010035

4. Welp A, Meier LL, Manser T. Emotional exhaustion and workload predict clinician-rated and objective patient safety. Frontiers in psychology 2014;5:1573. doi:10.3389/fpsyg.2014.01573 [published Online First: 2015/02/07]

5. Cimiotti JP, Aiken LH, Sloane DM, et al. Nurse staffing, burnout, and health care-associated infection. American journal of infection control 2012;40(6):486–90. doi:10.1016/j.ajic.2012.02.029 [published Online First: 2012/08/03]

6. Consiglio C. Interpersonal strain at work: A new burnout facet relevant for the health of hospital staff. Burnout Research 2014;1(2):69–75. doi:https://doi.org/10.1016/j.burn.2014.07.002

7. Leiter MP, Maslach C. Nurse turnover: the mediating role of burnout. Journal of nursing management 2009;17(3):331–9. doi:10.1111/j.1365-2834.2009.01004.x [published Online First:2009/05/12]

8. Shanafelt T, Sloan J, Satele D, et al. Why do surgeons consider leaving practice? Journal of the American College of Surgeons 2011;212(3):421–2. doi:10.1016/j.jamcollsurg.2010.11.006 [published Online First:2011/03/02]

9. Shanafelt TD, Dyrbye LN, West CP, et al. Potential Impact of Burnout on the US Physician Workforce. Mayo Clinic proceedings 2016;91(11):1667–68. doi:10.1016/j.mayocp.2016.08.016 [published Online First:2016/11/07]

10. Han S, Shanafelt TD, Sinsky CA, et al. Estimating the Attributable Cost of Physician Burnout in the United States. Ann Intern Med 2019;170(11):784–90. doi:10.7326/m18-1422 [published Online First:2019/05/28]

11. Lee PT, Loh J, Sng G, et al. Empathy and burnout: a study on residents from a Singapore institution. Singapore medical journal 2018;59(1):50–54. doi:10.11622/smedj.2017096 [published Online First:2017/10/13]

12. Tay WY, Earnest A, Tan SY, et al. Prevalence of Burnout among Nurses in a Community Hospital in Singapore: A Cross-Sectional Study. 2014;23(2):93–99. doi:10.1177/201010581402300202

13. Clough BA, March S, Leane S, et al. What prevents doctors from seeking help for stress and burnout? A mixed-methods investigation among metropolitan and regional-based australian doctors. Journal of clinical psychology 2019;75(3):418–32. doi:10.1002/jclp.22707 [published Online First:2018/11/16]

14. Guille C, Speller H, Laff R, et al. Utilization and barriers to mental health services among depressed medical interns: a prospective multisite study. Journal of graduate medical education 2010;2(2):210–4. doi:10.4300/jgme-d-09-00086.1 [published Online First:2010/06/01]

15. (ASAHP) AoSoAHP. What is Allied Health? 2015 [Available from: https://www.asahp.org/what-is.

16. K M. Jobs to Careers: Transforming the Front Lines of Health Care December 2012 [Available from: https://www.rwjf.org/en/library/research/2012/12/jobs-to-careers--transforming.

17. SingHealth. The Heartbeat of Healthcare: Allied Health Professionals; Many Talents, One Passion.

18. Welcome to the AHPC Singapore government agency website 2020 [Available from: https://www.healthprofessionals.gov.sg/ahpc.

19. Balogun JA, Titiloye V, Balogun A, et al. Prevalence and determinants of burnout among physical and occupational therapists. J Allied Health 2002;31(3):131–9. [published Online First:2002/09/14]

20. Gingras J, De Jonge LA, Purdy N. Prevalence of dietitian burnout. Journal of Human Nutrition and Dietetics 2010;23(3):238–43. doi:10.1111/j.13650-277X.2010.01062.x

21. Annual Report 2019: Allied Health Professions Council; 2019 [Available from: https://www.healthprofessionals.gov.sg/docs/librariesprovider5/forms-and-downloads/ahpc-annual-report-2019_final.pdf.

22. Ministry of Health S. National Health Survey 2010. In: Statistics R, ed., 2011.

23. Jackson CMSE. MBI: Human Services Survey for Medical Personnel. Maslach Burnout Inventory 2019

24. Rotenstein LS, Torre M, Ramos MA, et al. Prevalence of Burnout Among Physicians: A Systematic ReviewPrevalence of Burnout Among PhysiciansPrevalence of Burnout Among Physicians. JAMA 2018;320(11):1131–50. doi:10.1001/jama.2018.12777

25. Chao SF, McCallion P, Nickle T. Factorial validity and consistency of the Maslach Burnout Inventory among staff working with persons with intellectual disability and dementia. J Intellect Disabil Res 2011;55(5):529–36. doi:10.1111/j.1365-2788.2011.01413.x [published Online First:2011/03/23]

26. Maslach, Schaufeli, Leiter. Job burnout. Annual Review of Psychology 2001;52:397–422. doi:10.1146/annurev.psych.52.1.397

27. Dyrbye, West, Shanafelt. Defining burnout as a dichotomous variable. Journal of General Internal Medicine 2009;24:440. doi:10.1007/s11606-008-0876-6

28. Doulougeri K, Georganta K, Montgomery A. “Diagnosing” burnout among healthcare professionals: Can we find consensus? Cogent Medicine 2016;3(1) doi:10.1080/2331205X.2016.1237605

29. McCallister DE, Hamilton T. Transforming the Heart of Practice: An Organizational and Personal Approach to Physician Wellbeing: Springer International Publishing 2019.

30. Leiter MP, Maslach C. SIX AREAS OF WORKLIFE: A MODEL OF THE ORGANIZATIONAL CONTEXT OF BURNOUT. Journal of Health and Human Services Administration 1999;21(4):472–89.

31. Leiter M, Maslach C. Areas of Worklife: A Structured Approach to Organizational Predictors of Job Burnout2004:91–134.

32. Nguyen HTT, Kitaoka K, Sukigara M, et al. Burnout Study of Clinical Nurses in Vietnam: Development of Job Burnout Model Based on Leiter and Maslach’s Theory. Asian Nurs Res (Korean Soc Nurs Sci) 2018;12(1):42–49. doi:10.1016/j.anr.2018.01.003 [published Online First: 2018/02/22]

33. Jameson BE, Bowen F. Use of the Worklife and Levels of Burnout Surveys to Assess the School Nurse Work Environment. The Journal of School Nursing 2018:1059840518813697. doi:10.1177/1059840518813697

34. Lourel M, Gueguen N. [A meta-analysis of job burnout using the MBI scale]. Encephale 2007;33(6):947–53. doi:10.1016/j.encep.2006.10.001 [published Online First:2008/09/16]

35. Gascón S, Leiter MP, Stright N, et al. A factor confirmation and convergent validity of the “areas of worklife scale” (AWS) to Spanish translation. Health Qual Life Outcomes 2013;11:63–63. doi:10.1186/1477-7525-11-63

36. Mohr DC, Ho J, Duffecy J, et al. Perceived barriers to psychological treatments and their relationship to depression. Journal of clinical psychology 2010;66(4):394–409. doi:10.1002/jclp.20659

37. Sahlqvist S, Song Y, Bull F, et al. Effect of questionnaire length, personalisation and reminder type on response rate to a complex postal survey: randomised controlled trial. BMC Med Res Methodol 2011;11:62–62. doi:10.1186/1471-2288-11-62

38. Linden DVD, Keijsers GPJ, Eling P, et al. Work stress and attentional difficulties: An initial study on burnout and cognitive failures. Work & Stress 2005;19(1):23–36. doi:10.1080/02678370500065275

39. Yao Y, Yao W, Wang W, et al. Investigation of risk factors of psychological acceptance and burnout syndrome among nurses in China. Int J Nurs Pract 2013;19(5):530–8. doi:10.1111/ijn.12103 [published Online First:2013/10/08]

40. Kang K, Absher R, Granko RP. Evaluation of burnout among hospital and health-system pharmacists in North Carolina. American Journal of Health-System Pharmacy 2020;77(6):441–48. doi:10.1093/ajhp/zxz339

41. Silva SCPS, Nunes MAP, Santana VR, et al. Burnout syndrome in professionals of the primary healthcare network in Aracaju, Brazil. Cien Saude Colet 2015;20(10):3011–20. doi:10.1590/1413-812320152010.19912014

42. Zarei E, Ahmadi F, Sial MS, et al. Prevalence of Burnout among Primary Health Care Staff and Its Predictors: A Study in Iran. Int J Environ Res Public Health 2019;16(12) doi:10.3390/ijerph16122249 [published Online First:2019/06/28]

43. Yu H, Jiang A, Shen J. Prevalence and predictors of compassion fatigue, burnout and compassion satisfaction among oncology nurses: A cross-sectional survey. Int J Nurs Stud 2016;57:28–38. doi:10.1016/j.ijnurstu.2016.01.012 [published Online First: 2016/04/06]

44. Mason VM, Leslie G, Clark K, et al. Compassion fatigue, moral distress, and work engagement in surgical intensive care unit trauma nurses: a pilot study. Dimens Crit Care Nurse 2014;33(4):215–25. doi:10.1097/dcc.0000000000000056 [published Online First:2014/06/05]

45. Salyers MP, Bonfils KA, Luther L, et al. The Relationship Between Professional Burnout and Quality and Safety in Healthcare: A Meta-Analysis. J Gen Intern Med 2017;32(4):475–82. doi:10.1007/s11606-016-3886-9 [published Online First:2016/10/28]

46. Navarro-González D, Ayechu-Díaz A, Huarte-Labiano I. [Prevalence of burnout syndrome and its associated factors in Primary Care staff]. Semergen 2015;41(4):191–8. doi:10.1016/j.semerg.2014.03.008 [published Online First:2014/05/27]

47. Dewa CS, Jacobs P, Thanh NX, et al. An estimate of the cost of burnout on early retirement and reduction in clinical hours of practicing physicians in Canada. BMC Health Serv Res 2014;14:254. doi:10.1186/1472-6963-14-254 [published Online First:2014/06/15]

48. West CP, Dyrbye LN, Shanafelt TD. Physician burnout: contributors, consequences and solutions. Journal of Internal Medicine 2018;283(6):516–29. doi:10.1111/joim.12752

49. Brand SL, Thompson Coon J, Fleming LE, et al. Whole-system approaches to improving the health and wellbeing of healthcare workers: A systematic review. PLoS One 2017;12(12):e0188418. doi:10.1371/journal.pone.0188418 [published Online First:2017/12/05]

50. Ryan RM, Lynch MF, Vansteenkiste M, et al. Motivation and Autonomy in Counseling, Psychotherapy, and Behavior Change: A Look at Theory and Practice 1ψ7. The Counseling Psychologist 2010;39(2):193–260. doi:10.1177/0011000009359313

51. Hoi SY, Ismail N, Ong LC, et al. Determining nurse staffing needs: the workload intensity measurement system. Journal of Nursing Management 2010;18(1):44–53. doi:10.1111/j.1365-2834.2009.01045.x

52. Elbarazi I, Loney T, Yousef S, et al. Prevalence of and factors associated with burnout among health care professionals in Arab countries: a systematic review. BMC Health Serv Res 2017;17(1):491. doi:10.1186/s12913-017-2319-8 [published Online First:2017/07/19]

53. Ma LK. Medscape National Physician Burnout, Depression & Suicide Report 2019: Medscape Business of Medicine, 2019:29.

54. Bakker AB, Van der Zee KI, Lewig KA, et al. The relationship between the Big Five personality factors and burnout: a study among volunteer counselors. J Soc Psychol 2006;146(1):31–50. doi:10.3200/socp.146.1.31-50 [published Online First:2006/02/17]

